# Predicting pulmonary function from the analysis of voice: a machine learning approach

**DOI:** 10.1101/2021.05.11.21256997

**Authors:** Md. Zahangir Alam, Albino Simonetti, Rafaelle Billantino, Nick Tayler, Chris Grainge, Pandula Siribaddana, S. A. Reza Nouraei, James Batchelor, M. Sohel Rahman, Eliane V. Mancuzo, John W Holloway, Judith A. Holloway, Faisal I Rezwan

## Abstract

Providing proper timely treatment of asthma, self-monitoring can play a vital role in disease control. Existing methods (such as peak flow meter, smart spirometer) requires special equipment and are not always used by the patient. Using voice recording as surrogate measures of lung function can be used to assess asthma, which has good potential to self-monitor asthma and could be integrated into telehealth platforms. This study aims to apply machine learning approach to predict lung functions from recorded voice for asthma patients.

A threshold-based mechanism was designed to separate speech and breathing from recordings (323 recordings from 26 participants) and features extracted from these were combined with biological attributes and lung function (percentage predicted forced expiratory volume in 1 second, FEV_1_%). Three predictive models were developed: (a) regression models to predict lung function, (b) multi-class classification models to predict the severity, and (c) binary classification models to predict abnormality. Random Forest (RF), Support Vector Machine (SVM), and Linear Regression (LR) algorithms were implemented to develop these predictive models. Training and test samples were separated (70%:30% using balanced portioning). Features were normalised and 10-fold cross-validation used to measure the model’s training performances on the training samples. Models were then run on the test samples to measure the final performances.

The RF based regression model performed better with lowest root mean square error = 10.86, and mean absolute score = 11.47, as compared to other models. In predicting the severity of lung function, the SVM based model performed better with 73.20% accuracy. The RF based model performed better in binary classification models for predicting abnormality of lung function (accuracy = 0.85, F_1_-score = 0.84, and area under the receiver operating characteristic curve = 0.88).

The proposed machine learning approach can predict lung function (in terms of FEV_1_%), from the recorded voice files, better than other published approaches. These models can be extended to predict both the severity and abnormality of lung function with reasonable accuracies. This technique could be used to develop future telehealth solutions including smartphone-based applications which have potential to aid decision making and self-monitoring in asthma.

## 1. Introduction

Asthma is a common respiratory condition that affects 235 million people worldwide.^1^ Around 5.4 million people in the UK are currently receiving treatment for asthma, approximately 1 in 11 children and 1 in 12 adults.^2^ Every 10 seconds, at least one person is facing a potentially life-threatening asthma attack in the UK, and on an average, three people die from it daily, regardless the effective treatments developed in recent years.^3^ Appropriate, effective management and treatment for asthma is therefore of vital importance.

Many different techniques can monitor the complex nature of asthma, including subjective symptom assessments, lung function testing, and measurement of biomarkers. Regular monitoring of asthma can help patients receive appropriate treatment in time, which can help to reduce symptoms, frequency of exacerbation, and risks of hospitalisation. The ability to monitor asthma and modify treatment appropriately could help to reduce both disease morbidity and the economic cost of treatment. Identifying symptoms via questionnaire and lung function measurement via spirometry identifying of biomarkers (e.g. exhaled nitric oxide or sputum eosinophils) can all be used in regular monitoring of asthma.^4^ In practice, however, the combination of these is impractical in community-based care due to expense and/or complexity.

Self-monitoring of asthma has the potential to play an important role in empowering the patient and maintaining disease control; such monitoring needs to be simple, convenient, and accurate. Equipment such as smart spirometers and, accompanying smartphone apps used to record peak expiratory flow rates (PEFR) and provide reminders to manage asthma more efficiently are currently available to simplify self-monitoring.^5^ However, smart spirometers are still expensive for personal use. As more people use smartphones, an application measuring lung function that could alert patients to modify their treatment without the need for a spirometer would be a convenient and inexpensive way to monitor asthma, particularly in Lower and Middle Income Country (LIMC) settings.

At present, assessment of the ability to speak and the sounds associated with breathing are a recognised part of an assessment of asthma severity, such as: “speaking full sentences” to “unable to speak at all” together with wheeze on auscultation.^6–8^ Although no standardised assessment or quantitative measures of these features have been developed, the effects on speech and breathing patterns and sounds due to increased airway resistance are noticeable in acute asthma.^9^ Thus, pitch from speech and quality of the breathing sound can potentially be utilised as surrogate measures of symptoms and/or to predict lung function, which can then be used to monitor asthma.

Three kinds of sounds have been analysed to predict lung function using machine learning techniques: (1) lung and breathing sounds from the chest, (2) symptom-based sounds (such as a cough sound), and (3) voice sounds. Quantitative breath sound measurements, such as Vibration Response Imaging (VRI), have been used to predict postoperative lung function.^10,11^ Cough and wheeze sound-based analyses have been shown to have potential in predicting spirometer readings.^12–14^

In parallel to symptom-based sounds (such as a cough sound), there are a number of studies, which involve voice sounds only. A recent review identified 20 studies to date. It confirmed the idea of respiratory function correlating significantly to phonation sound. Some of these studies showed that voice evaluation might allow recognition of asthma contributing to voice dysfunction subjected to lung function.^15^ However, most of these studies required the use of specialised instruments and software to quantify specialised phonetic sounds. Using machine learning techniques, one of these studies showed that sustained phonation of the vowel sound demonstrated potential utility in the diagnosis and classification of severity of asthma.^16^

Assessing the quality of sound produced by an asthma patient, primarily via speech, is a common way to assess acute asthma. We have previously demonstrated that recorded speech correlates well with lung function during induced bronchoconstriction.^17^ To date, only two studies have utilised machine learning techniques to predict lung function from the recorded voice. Saleheen et al. proposed a convenient mobile-based approach that utilises a monosyllabic voice segment called A-vowel’ sound or ‘Aaaa…’ sound from voice to estimate lung function.^18^ Chun et. al. proposed two algorithms for passive assessment of pulmonary conditions: one for detection of obstructive pulmonary disease and the other for estimation of the pulmonary function in terms of ratio of forced expiratory volume in 1 second (FEV_1_) and forced vital capacity (FVC) also denoted as FEV_1_/FVC and percentage predicted FEV_1_ (FEV_1_%).^19^ However, these studies showed moderate performance and did include comparison with previous studies.

This study proposes a new methodology to predict lung function from recorded speech using machine learning techniques to monitor asthma. Bronchoprovocation tests were given to participants to help diagnose asthma, and their voices were recorded for 1 min while the subjects read standard texts with lung function measured. This study aims to identify features from recorded speech files that correlate with measured lung function. We subsequently use those features to predict lung function, potentially enabling identification of deterioration of asthma control via a smartphone application in the future.

## 2. Methods

### 2.1. Dataset

Twenty-six non-smoking, clinically stable subjects, with physician-diagnosed mild atopic asthma, were recruited on step one of treatment according to 2012 GINA guidelines.^20^ The study was approved by the local ethics committee (number 12/EE/0545), by the Medicines and Healthcare Products Regulation Agency (MHRA) (MHRA number 11709/0246/001-0001).^21^ The study was performed in compliance with the protocol and additional methodologic details provided in the supplementary material. All participants underwent a standardised inhaled methacholine challenge^21^ and after each challenge dose, the participant read a standardised text for 30s into a digital recorder fitted with an external microphone set at 10 cm from the mouth (Olympus DM450 Speech Recorder with Olympus ME34 Microphone, Tokyo, Japan).. After each dose of the bronchial challenge, the voice of each subject was recorded, and lung function was measured as FEV_1_% predicted. Spirometry was performed with a dry bellows spirometer (Vitalograph, UK) and the best of at least three successive readings within 100 ml of each other was recorded as the FEV_1_ in accordance with established guidelines.^22^ In total 323 voice recorded sound files with their associated FEV_1_% were recorded for these 26 subjects. Details of the method is shown in the Supplementary Methods section.

### 2.2. Separation of breathing and speech segments from sound files

An exploratory analysis was carried out on a segment of speech and breathing separately in the frequency domain and considerable differences were noticed in the spectrograms generated by librosa^23^ (Supplementary Figure S1). The parts of the sound file containing breathing and speech were separated from five randomly selected sound files using Audacity software.^24^ Features (including roll-off at 85%/95%, spectral fitness, root mean square energy, zero crossing rate, spectral centroid, spectral bandwidth, spectral contrast, spectral flatness, mean amplitude, and mean breath cycle duration) (described in Supplementary Methods) were extracted for individual breathing and speech segments, using the librosa tool. These features were analysed to explore differences between breathing and speech segments and determine the appropriate thresholds to separate breathing and speech segments.

### 2.3. Feature extraction

After analysing the values for individual speech and breathe segments, only five features (Spectral contrast, Roll-off at 95%, Root mean squared energy, Spectral bandwidth, and Mean amplitude) showed substantial differences between breathing and speech segments (Supplementary Figure S2). Based on the observed information, using a threshold, these 5 features were defined (Supplementary Table S1), which separate the breathing and speech segments from all available sound files.

All extracted features were Min-max normalised. As there was a low number of features, it was impossible to utilise a feature engineering method to identify informative features. The use of Pearson correlation coefficient calculated the correlation between the features and FEV_1_%.

### 2.4. Predictive model development

Training and testing samples were separated randomly at a ratio of 70%:30%, respectively (i.e., the training dataset contained 70% of the samples, whereas the testing dataset kept the remaining 30% of the samples). This defined the following three types of predictive models:

Model_1_: A regression model to predict FEV_1_% predicted based on the features extracted from recorded sound data. The techniques and the feature set for which this model performs best were applied for the other following models. The performances of these models are reported in terms of Root Mean Square Error (RMSE) and mean absolute error (MAE).

Model_2_: A multi-class classification model to predict the severity of abnormality of lung function according to American Thoracic Society (ATS) grades (as defined in Supplementary Table S2).^25^

Model_3_: A binary classification model to predict FEV_1_% classified either as normal or abnormal based on ATS grades for the severity of abnormal lung function (Supplementary Table S2), where lung function is normal if FEV_1_% > 80%, otherwise lung function is abnormal.

Three machine learning algorithms, including Random Forest (RF), Support Vector Machine (SVM) (using Radial Basis Function kernel), and Linear Regression (LR), were implemented to develop the predictive models. Training the models was undertaken on the “training set”, and 10-fold cross-validation was used to measure the models training performances. The Models use Default values of hyperparameters, and tuning did not show any improvements over default parameters. Finally, the models were run on the testing samples to assess the final performances.

Further basic biological attributes, including sex, height and weight of the subjects were added as features in addition to the features extracted from the sound file as mentioned above to develop three additional models (denoted as Model_1P_, Model_2P_, and Model_3P,_ respectively for Model_1_, Model_2_, and Model_3_).

Initially, we investigated the effect of features extracted from speech and breathing parts individually and combined on lung function using Model1. We also explored the problem with imbalances of distribution of lung function in random partitioning and performed balanced partitioning of training and test samples as follows:

1. Using intervals of 5% on the FEV1% values 15 groups were prepared. For example, groups are 51-55%, 56-60%, 61-65% etc.
2. Based on the groups sample distribution was prepared.
3. Based on the sample distribution in each group balanced training and test sets were prepared, such that training and test sets followed the same distribution.

## 3. Results

Table 1 showed the severity of abnormal lung function among 323 data samples. It is evident that 72.14% of samples exhibited normal lung function during corresponding recording and the rest (27.86%) exhibited abnormal lung function.

**Table 1:** Percentage of the samples in terms of severity of abnormality of the lung function

| Severity of lung function | Percentage among samples |
| --- | --- |
| Normal | 72.14 % |
| Mildly abnormal | 15.78 % |
| Moderately abnormal | 9.29 % |
| Moderate to severely abnormal | 2.79 % |

### 3.1. Feature extraction from recorded voice files

Fourteen breathing segments and nine speech segments were retrospectively extracted from the sound files. (Supplementary Table S3). Results show no correlation between the features and FEV_1_% (Supplementary Figure S3).

### 3.2. Lung function prediction in terms of FEV1% (Model1)

#### 3.2.1. Effect of speech and breathing features on prediction

Initially, we explored the ability to predict lung function from extracted features of speech and breath both individually and in combination.,. Regression models developed using the combined features from speech and breathing to predict FEV_1_%, showed lower mean absolute error (MAE) than that of models developed from features from speech and breathing separately (Figure 1A). The RF model (Model_1(RF)_) performed better in comparison to all other algorithms (the lowest Root Mean Square Error, RMSE = 12.59) (Figure 1B).

**Figure 1:**
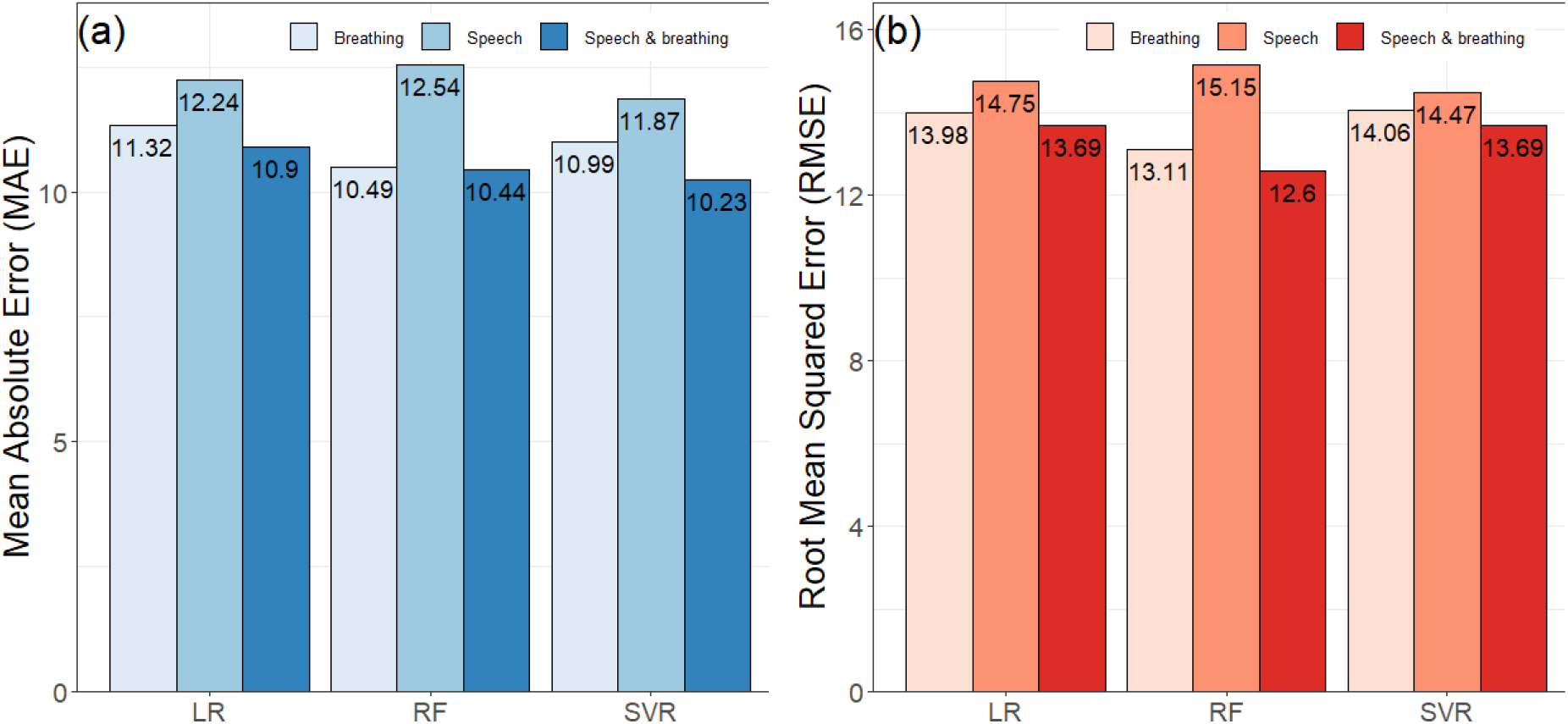
Impact of speech and breathing features individually and combinedly on model development. (a) shows the performances of the models in terms of mean absolute error (MAE) and (b) presents the performances of the models in terms of root mean squared error (RMSE). Here, LR = Linear Regression, RF = Random Forest, and SVR = Support Vector Regression.

#### 3.2.2. Effect of balanced partitioning of the training and the testing sets

The samples were not uniformly distributed amongst the ranges of FEV_1_% (Figure 2). Frequency of the samples is the highest around 100 of the FEV_1_% values and no sample was found with the FEV_1_% ≤ 50. As a result, samples were not uniformly distributed among the ranges of FEV1% values. Therefore, when the training and test samples were divided randomly, the pattern in the training dataset may not follow the pattern in the test dataset (Supplementary Figure S4A). The balanced separation of training and test samples shows a similar pattern of the samples among each range of FEV_1_% (Supplementary Figure S4B).

**Figure 2:**
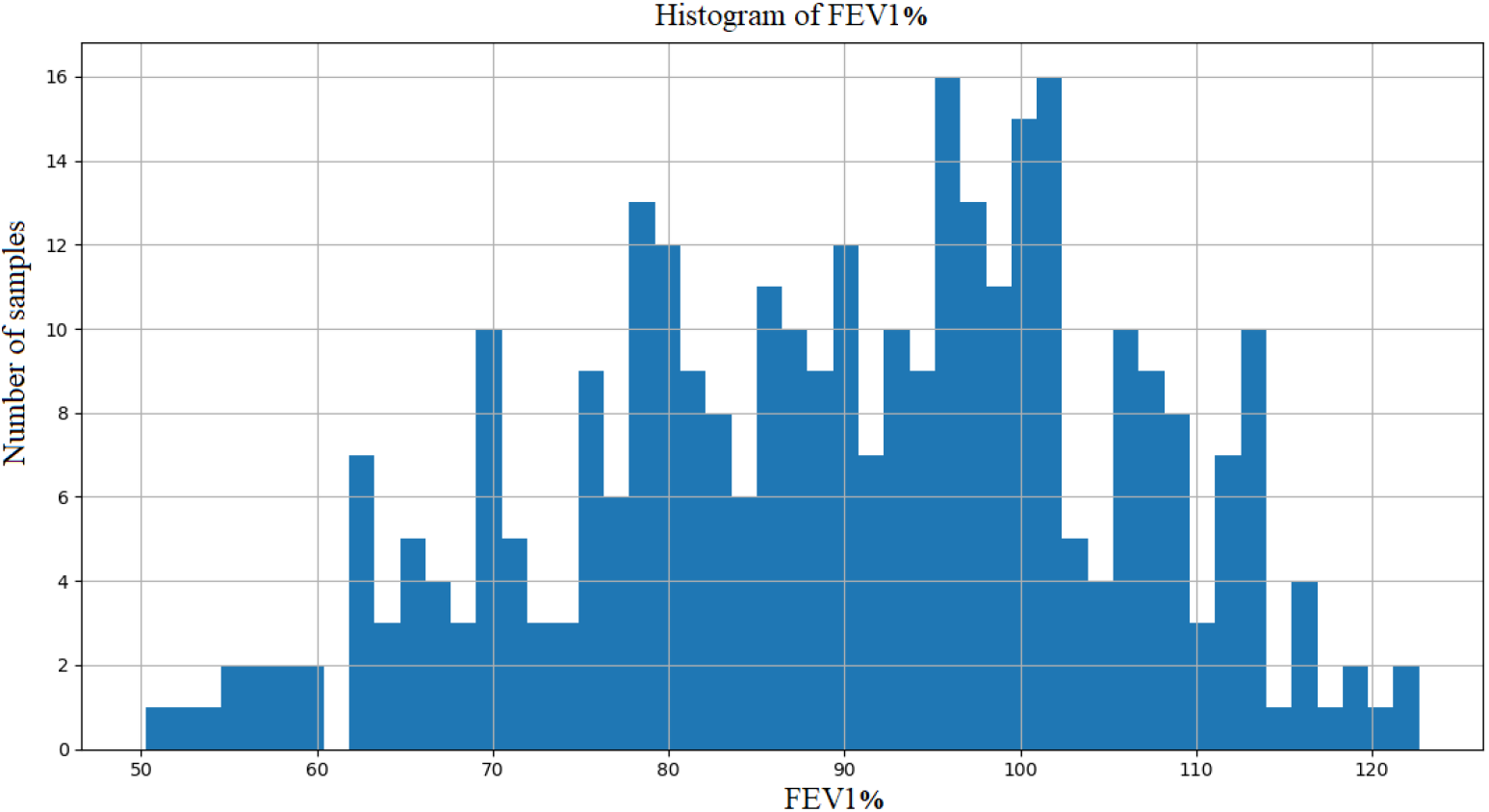
Histogram of FEV_1_% values of the samples. It represents the frequency distribution of FEV_1_% values of the samples. FEV_1_% values are represented in x-axis and frequency of the samples per FEV_1_% is shown in y-axis.

Balanced partitioning of the training and the test sets led to improved performance compared to random partitioning. This is evident for all regression models, where balanced partitioning shows lower RMSE and MAE scores in comparison to the random partition model (Figure 3). Again, the RF based model (Model_1(RF)_,) performed better than other models (RMSE = 12.51 and MAE = 9.83).

**Figure 3:**
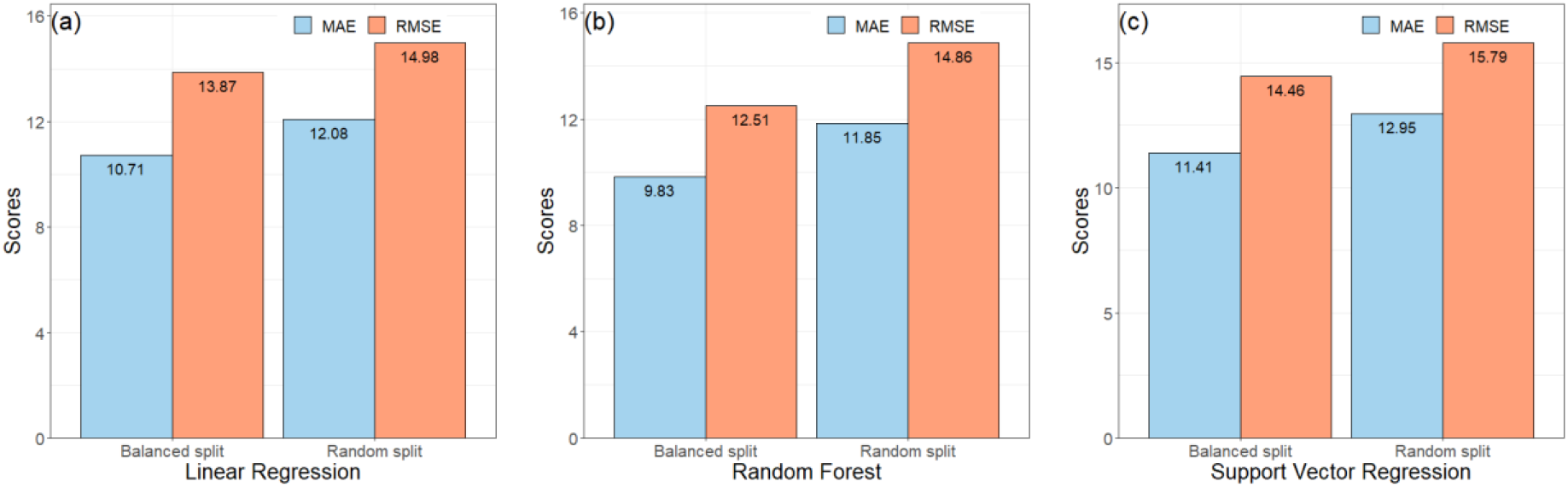
Effects of the balanced partitioning of the training and testing sets. The performances of the Model_1(LR)_, Model_1(RF)_ and Model_1(SVM),_ with and without balanced partitioning, are shown in (a), (b) and (c) respectively here. Balanced split represents the training and the test sets were created based sample distribution shown in Table 1 and the random split represent training and the test sets were created based on random partitioning. Here, MAE = Mean Absolute Error and RMSE = Root Mean Squared Error.

#### 3.2.3. Effects of the phenotypes on the predictive models

The performance of the models when biological attributes were added are shown in Figure 4. The RF based algorithm performed better with MAE (%) score of 10.86 and RMSE score of 11.47 as compared to other algorithms. Table 2 shows the comparison of the Model_1(RF)_, and the Model_1P(RF)_) for predicting FEV1%. Model_1P(RF)_) showed better predictive performance than that of Model_1(RF)_).

**Table 2:** Performances of the Model_1_ (without sex, weight & height) and Model_1P_ (with sex, weight & height) based on Random Forest for predicting lung function in terms of FEV1% (regression)

| Models | MAE | RMSE |
| --- | --- | --- |
| Model<sub>1(RF)</sub> (without sex, weight & height) | 9.83 | 12.51 |
| Model<sub>1P(RF)</sub> (with sex, weight & height) | 8.96 | 11.48 |

**Figure 4:**
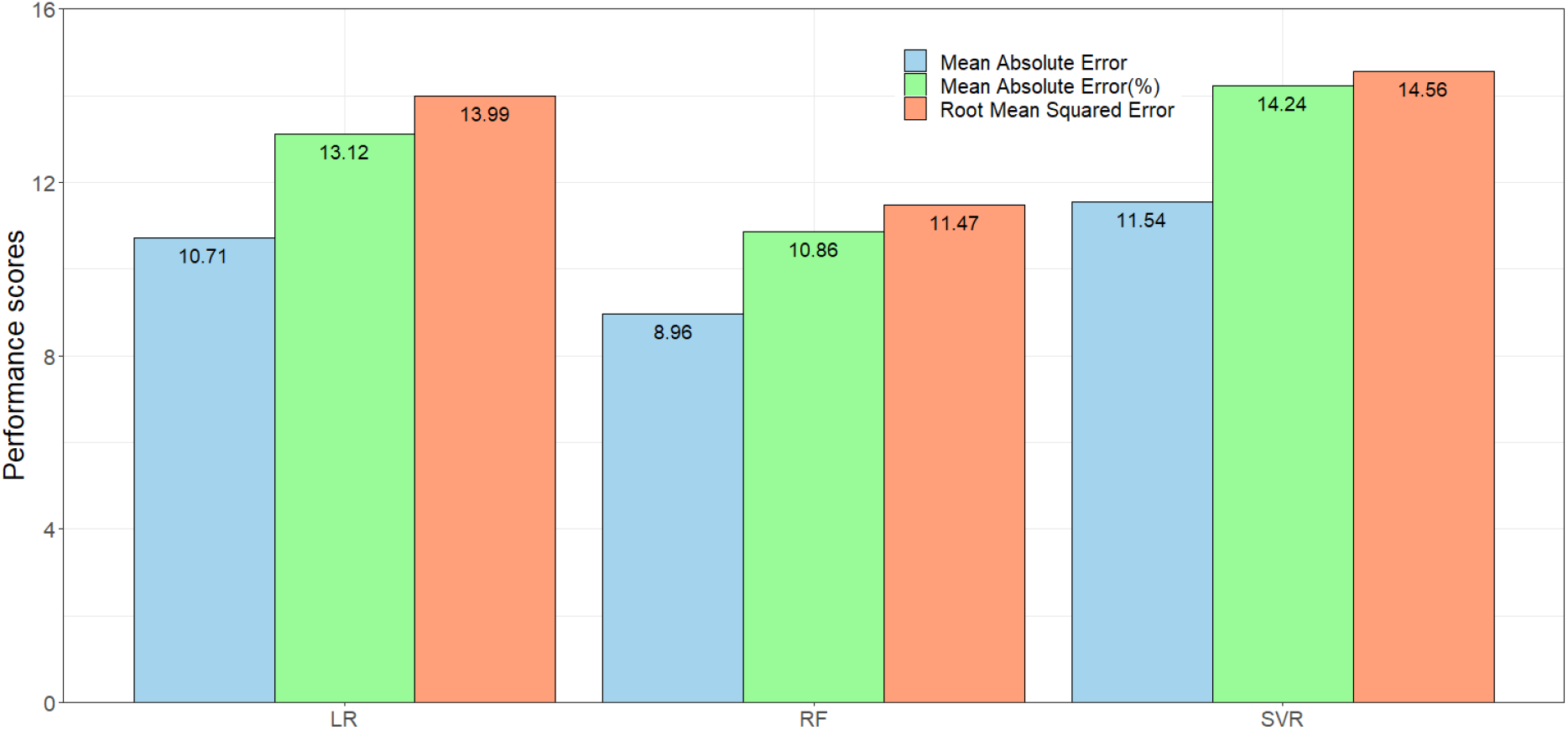
The performance of the regression models (Model_1P_ with sex, height and weight have been added with the features from the voice sounds). Here, LR = Linear Regression, RF = Random Forest, and SVR = Support Vector Regression

### 3.3. Severity of abnormality of lung function prediction (Model_2_)

The performance of Model_2_ and Model_2P_ in predicting lung function severity from the sound files, is shown in Table 3. Model_2(SVC)_ predicted abnormal lung function with 71% accuracy, while Model_2P(SVC)_ predicted this abnormality with 73.2% accuracy.

**Table 3:** Comparison of the performances (accuracy) of Model_2_ (without sex, weight & height) and Model_2P_ (with sex, weight & height) in predicting the severity of abnormality of lung function

| Algorithms | Model<sub>2</sub> | Model<sub>2P</sub> |
| --- | --- | --- |
| Linear Regression | 0.64 | 0.66 |
| Random Forest | 0.68 | 0.71 |
| Support Vector Classifier | 0.71 | 0.73 |

### 3.4. Normal vs. abnormal lung function prediction (Model_3_)

The performance of the models (without and with biological attributes) in predicting normal vs. abnormal lung function are shown in Tables 4 and 5 (detailed in Supplementary Tables S4 and S5). The best performance (without adding physical attributes) was observed for the RF model, Model_3(RF)_, with 80% accuracy and 79% F_1_-score. The RF based model clearly performs better (AUC = 0.84) than the other models (Figure 5A).

**Table 4:** Comparison of the performances of Model_3_ (without sex, weight & height) in predicting the abnormality of lung function. Here, RF = Random Forest, SVC = Support Vector Classifier and LR = Linear Regression

|  | Precision | Recall | F<sub>1</sub>-score | Accuracy | Sensitivity | Specificity | AUC |
| --- | --- | --- | --- | --- | --- | --- | --- |
| LR | 0.77 | 0.78 | 0.76 | 0.78 | 0.37 | 0.94 | 0.78 |
| RF | 0.80 | 0.80 | 0.79 | 0.80 | 0.44 | 0.94 | 0.84 |
| SVC | 0.80 | 0.78 | 0.74 | 0.78 | 0.26 | 0.99 | 0.78 |

**Table 5:** Comparison of the performances of Model_3P_ (with sex, weight & height) in predicting the abnormality of lung function. Here, RF = Random Forest, SVC = Support Vector Classifier and LR = Linear Regression

|  | Precision | Recall | F<sub>1</sub>-score | Accuracy | Sensitivity | Specificity | AUC |
| --- | --- | --- | --- | --- | --- | --- | --- |
| LR | 0.82 | 0.82 | 0.81 | 0.83 | 0.48 | 0.96 | 0.84 |
| RF | 0.84 | 0.85 | 0.84 | 0.85 | 0.63 | 0.93 | 0.88 |
| SVC | 0.81 | 0.80 | 0.78 | 0.80 | 0.37 | 0.97 | 0.79 |

**Figure 5:**
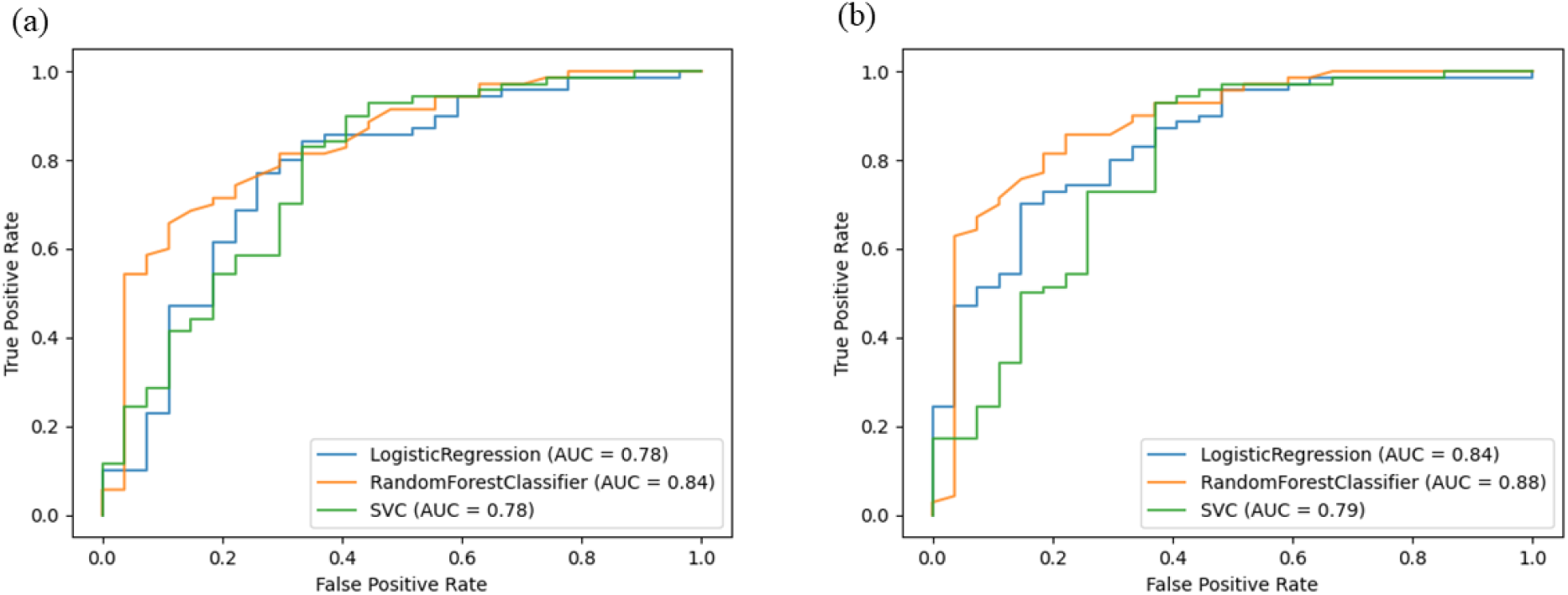
Receiver operating characteristic (ROC) curve plots of Model_3_ and Model_3P_. These plots show the area under Receiver Operating Characteristic curve of model’s showing performance for predicting normal vs. abnormal lung function. (a) showing the ROC curve for Model_3_ and (b) showing the ROC curve for Model_3p_.

This held true when adding physical attributes, with the RF-based model again showing the best performance (accuracy = 85%, F1-score = 84%, and AUC = 88% AUC (Table 5 and Figure 5B).

## 4. Discussion

This study focused on predicting lung function from recorded voice sounds in three ways and has developed a predictive model (for FEV_1_%), which can be applied in real-time applications for early intervention in asthma management. A model to predict the severity of abnormal lung function according to the ATS^23^ was also developed, as well as a model to predict normal vs abnormal lung function (i.e., FEV_1_% ≤ 80). By detecting abnormal lung function, this can prompt the patient to take appropriate action to manage their condition.

A threshold-based mechanism was defined to separate the breathing and speech features from the recorded sound files. A suitable threshold to separate the breathing and speech chunks was devised by utilising this mechanism, extracted 23 features to develop the predictive models. The results indicated that using the breathing and speech features in combination. They improved the performance of the predictive model’s performance model. This finding is consistent with standard clinical practice to identify acute asthma by listening to speech and breathing patterns.

Handling partitioning of the training and the testing dataset is an important factor in developing the prediction model. Considering the American Thoracic Society Grades for the severity of a Pulmonary Function Test Abnormality^26^, this study proposed a balanced partitioning technique for predicting FEV_1_% for asthma patients and, consequently, the model’s performance has improved comparing with random partitioning on handling imbalanced data.

Initially, the RF based predictive models showed better performance as compared to other models except in the prediction of severity of abnormality of the lung function. The RF based models predicted FEV_1_% with lower RMSE and MAE, and abnormality in lung function with high accuracy whereas SVM predicted severity of lung function with higher accuracy compared with that of RF and LR based models. The feature space in this study is not high dimensional (with only 26 features in total). The results are consistent with other studies that have reported a better performance of RF based models when working with a limited number of features.^14,19^ Generally, SVM is applied on highly dimensional space for best results. In addition, the correlational matrix showed no strong correlation of any feature with FEV_1_%, and RF performs better with non-linear problems.

Furthermore, due to their nature, RF models are less likely to overfit. While most of the scores of the RF-based models are quite reasonable, the sensitivity of the RF based classifier to predict abnormality of lung function was not high (sensitivity = 44%). This could be due to the irregular distribution of the samples to normal vs. abnormal lung function (approximately 3:1). Although RF based models (Model_1_ and Model_3_) showed better performance, the SVM based model performed well on predicting severity of lung function (Model_2_). This is possibly due to grouping samples (i.e., grouping of FEV_1_%) based on the severity of abnormality of the lung function and heterogeneous distribution of the samples into these groups (e.g., the samples with normal and with moderate to severely abnormal lung functions are 72.14% and 2.79% respectively in the dataset).

Adding biological attributes (sex, height and weight), to the model, along with the features extracted from speech and breathing, improved the performances of the models. This improvement was observed for all three methods (RF, SVM and LR) used in this study.

The predictive models used many pulmonary features, such as Mean Breath Cycle Duration, Breath Number, which relate to the airway resistance of patients with pulmonary disease (e.g. asthma). Breathing becomes more difficult for people with obstructive pulmonary disease due to increased airway resistance. As their pulmonary symptoms worsen, they frequently notice increased breathlessness and may have higher respiratory rates.^27^ Therefore, the pulmonary features extracted in this study are believed to have greater capability in predicting lung function. Similarly, other features, such as Roll off 95%, Mean Amplitude, Spectral Bandwidth etc., are also believed to be important in prediction tasks in this context. An earlier study reported a higher rate of increase in the intensity of the sound for equal increments in flow rate in chronic bronchitis and asthma than in healthy subjects.^28^ The inclusion of these pulmonary features is an essential attribute of this research in this context.

Only two recent studies have used voice sounds to predict lung function. Saleheen et al. extracted the “A-vowel” segments from the voice sound and then extracted features from the ‘A-vowel’ sounds and predicted lung function in terms of the FEV_1_/FVC ratio.^18^ Due to the lack of the FVC values in this study, the results are not directly comparable. Chun et al. developed models to predict lung function in terms of the FEV_1_/FVC ratio and FEV_1_%.^19^ Their reported prediction efficiency in terms of MAE (%) score is 20.6%, which is quite large for any regression problem. The RF based regression model reported in this research work achieved a MAE (%) score of 10.86% which shows significant improvement over that of Chun et al.

This pilot study has limitations, including the limited sample size and the range of machine learning algorithms utilised to develop the predictive models. To overcome this, the balanced partitioning technique was applied. The performance of the predictive regression model in estimating FEV1% values is reasonable, and this study presents a better performance compared with previous studies found in the literature, in addition to being able to predict normal vs abnormal lung function and the severity of abnormality of lung function. Considering the generalization of the model to avoid overfitting issue, 10-fold cross validation was applied during training of the model. Furthermore, no feature selection method was applied to identify important features among the 23 features. However, due to the limited number of features, this study did not consider feature engineering

Future work with more samples and using an external dataset will allow the predictive model to be better generalised and allow validation. A recent study included Mel Frequency Cepstral Coefficient (MFCC) value as a feature to predict COVID-19 subjects from a forced-cough cell-phone recording.^29^ In contrast, our study has used 23 features excluding MFCC due to the nature (breathing and speech) of pattern-finding for acute asthma prediction from sound files. MFCC represents a full signal at a time in the signal processing. On the other hand, 23 features extracted here present the micro-information of different parts of a signal (e.g., breathing chunks and speech chunks of a voice sound file). However, the application of MFCC along with these features may have the potential to improve the predictions.

## 5. Conclusion

Asthma puts pressure on health services due to the associated cost and workforce required to treat and care for the people with the condition. Therefore, regular monitoring and early intervention can help control the disease, reducing hospital admissions and therefore cost. The predictive models developed in this study can be implemented in smartphone applications offering a convenient and straightforward way to predict lung function. Embedding the algorithm in an app for self-monitoring asthma will potentially enable patients to achieve improved symptom control, availing early and appropriate medication and reducing costs.

## Data Availability

Access to data and source code can be made available upon request.

## Contributors

All authors reviewed the literature, read and approved the final Article. JAH and FIR conceptualised the study. JWH, JAH and FIR designed the study. NT and CG collected the data. MZA, AS and RB analysed data. ZAH, JAH and FIR contributed to data interpretation and drafted the Article. NT, CG, PS, SARN, JB, MSR, EVM and JWH critically reviewed the Article.

## Date sharing

Access to data and source code can be made available upon request.

## Acknowledgements

The authors thank the Southampton Biomedical Research Unit nurses, all the patients and health care professionals for their participation.

