## Supplementary materials for "Predicting pulmonary function from the analysis of voice: a machine learning approach"

**Supplementary Method:**

**Allergen challenge:**

An allergen challenge test involves directly exposing a person to a substance to try to find out whether the person is allergic to it.

**Bronchial challenge**

A bronchial challenge test is a medical test used to assist in the diagnosis of asthma. The person's normal lung function will be measured first (also known as his/her baseline breathing). Then he/she will start the test. During the test, he/she will be asked to inhale doses of methacholine, a drug that can cause narrowing of the airways. If his/her lung function drops by 20% or more from his/her baseline, he/she may have asthma. The test takes about 90 minutes. The breathing test will be repeated after each dose of methacholine to measure the degree of narrowing or constriction of the airways, therefore, multiple test results will be produced from a person.

**Forced Expiratory Volume (FEV)**

Forced expiratory volume (FEV) measures how much air a person can exhale during a forced breath. The amount of air exhaled may be measured during the first second ($FEV_1$), second seconds ($FEV_2$), and/or third seconds ($FEV_3$) of the forced breath. A lower than normal FEV1 reading suggests that the person may be experiencing a breathing obstruction. In other words, a person who has asthma or COPD has a lower $FEV_1$ result than a healthy person. The normal values for FEV1 vary from person to person. They are based on standards for an average healthy person of their age, race, height, and gender. Each person has their own predicted FEV1 value based on these factors mentioned above. We can get our predicted normal value with a spirometry calculator. If we know our $FEV_1$ value already, we can enter it as well, and the calculator will tell us what percent of the predicted normal value our result is.

$FEV_1$ can be converted to a percentage of the predicted normal value, termed as $FEV_1\%$. It is defined as follows:

$$\mathrm{FEV_1\%} = \left(\frac{\mathrm{FEV_1}}{\mathrm{Predicted\ FEV_1}}\right) * 100$$

**Reading material for participants.**

Instructions: please read the following text at the same volume as if you were talking on the telephone. If you reach the end of the text then please start again from the beginning.

When we got to the strange house it began to snow in quite a different way. A mass of tired old clouds opened and flung snow at us, all of a sudden and just anyhow. They weren’t ordinary snowflakes – they fell straight down in large sticky lumps, they clung to each other and sank quickly and they weren’t white, but grey. The whole world was as heavy as lead. Mummy carried in the suitcases and stamped her feet on the doormat and talked the whole time because she thought the whole thing was such fun and that everything was different. But I said nothing because I didn’t like this strange house. I stood in the window and watched the snow falling, and it was all wrong. It wasn’t the same as in town. There it blows black and white over the roof or falls gently as if from heaven, and forms beautiful arch over the sitting-room window. The landscape looked dangerous too. It was bare and open and swallowed up the snow, and the trees stood in black rows that ended in nothing. At the edge of the world there was a narrow fringe of forest. Everything was wrong. It should be winter in town and summer in the country. Everything was topsy-turvy. The house was big and empty, and there were too many rooms. Everything was very clean and you could never hear your own steps as you walked because the carpets were so big and they were as soft as fur.

(Source: Tove Jansson (2006). *A winter book.* London: Sort Of Books.)

**Details of the features**

1. **Roll-off at 85%/95%:** It represents the value in frequency below which is possible to find the 85%/95% of the total frequencies.
2. **Spectral flatness:** It is a measure to quantify how much noise-like a sound is, as opposed to being tone-like.
3. **Root mean squared energy:** The energy of a signal corresponds to the total magnitude of the signal. For audio signals, that roughly corresponds to how loud the signal is. The energy in a signal is defined as:

   $$\boldsymbol{Energy} = \sum_{\mathbf{1}}^{\boldsymbol{N}} |\boldsymbol{x}(\boldsymbol{n})|^{\mathbf{2}}$$

   where x[1] is a frame of a signal and n is the length of a frame.
   The root mean squared energy in a signal is defined as:

   $$\sqrt{\frac{\mathbf{1}}{\boldsymbol{N}} \sum_{\mathbf{1}}^{\boldsymbol{N}} |\boldsymbol{x}(\boldsymbol{n})|^{\mathbf{2}}}$$

4. **Zero Crossing Rate:** It indicates the number of times that a signal crosses the horizontal axis.
5. **Spectral centroid:** It is a measure used in digital signal processing to characterize a spectrum. It indicates where the centre of mass of the spectrum is located. Perceptually, it has a robust connection with the impression of brightness of a sound.
6. **Spectral bandwidth:** It is a measure of the extent of the spectrum. It is the wavelength interval in which a radiated spectral quantity is not less than half of its maximum value.
7. **Spectral contrast:** It considers the strength of spectral peaks and spectral valleys in each sub-band separately.
8. **Spectral Flatness:** It is defined as the ratio of the geometric mean to the arithmetic mean of a power spectrum.
9. **Mean amplitude:** It is the mean amplitude of various frequencies present in a sound signal. It is measured using Short Term Fourier transformation (STFT) of a signal. STFT converts a signal in such a way so that we can understand the amplitude of given frequency at a given time.
10. **Mean breath cycle duration:** Breath cycle duration is the duration from one breath chunk to the next breath chunk. The mean breath cycle duration is the mean of all breath cycle durations.

**Supplementary Figures:**

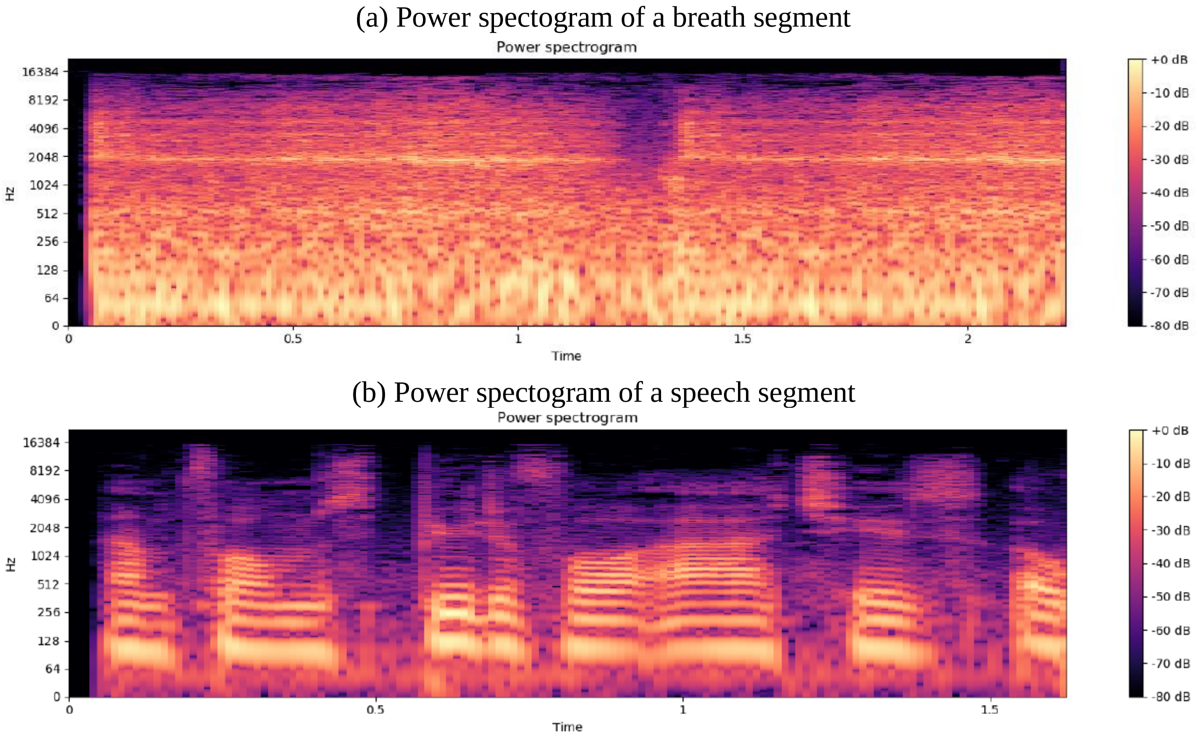


Supplementary Figure S1: Power spectrograms of a breathing chunk, and of a speech chunk from a sound file. It presents two power spectrograms: [a] Power spectrogram of a breathing chunk, [b] Power spectrogram of a chunk of a speech. In these spectrograms, we can observe that the breath segment exhibits higher frequencies, and the power of the signal is distributed more evenly throughout the time interval.

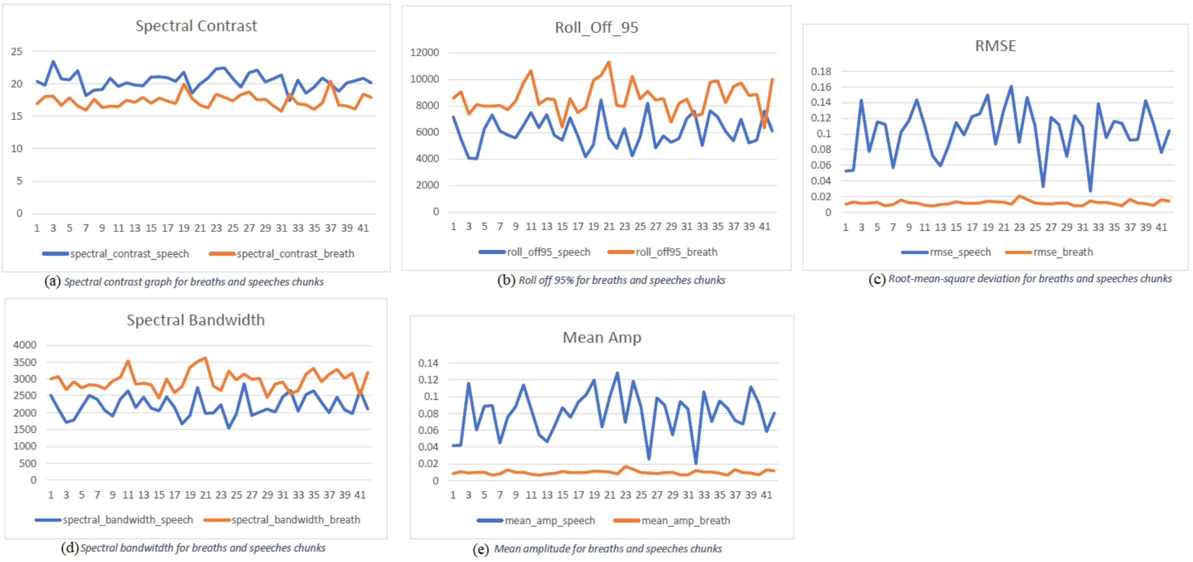


Supplementary Figure S2: Comparison of the breathing and speech chunks. The breathing chunks are compared with the chunks of the speeches. The features (please refer to Supplementary Background 1.5 for the details) have been extracted (by python package Librosa) from these chunks separately. Only 5 of them have shown substantial differences in terms of the features' values between the breathing chunks and the speech chunks. They are as follows: [a] Spectral contrast graph for the breathing chunks and the speech chunks; [b] Roll off at 95% for for the breathing chunks and the speech chunks; [c] Root mean squared energy deviation for the breathing chunks and the speech chunks; [d] Spectral bandwidth for the breathing chunks and the speech chunks; [e] Mean amplitude for the breathing chunks and the speech chunks.

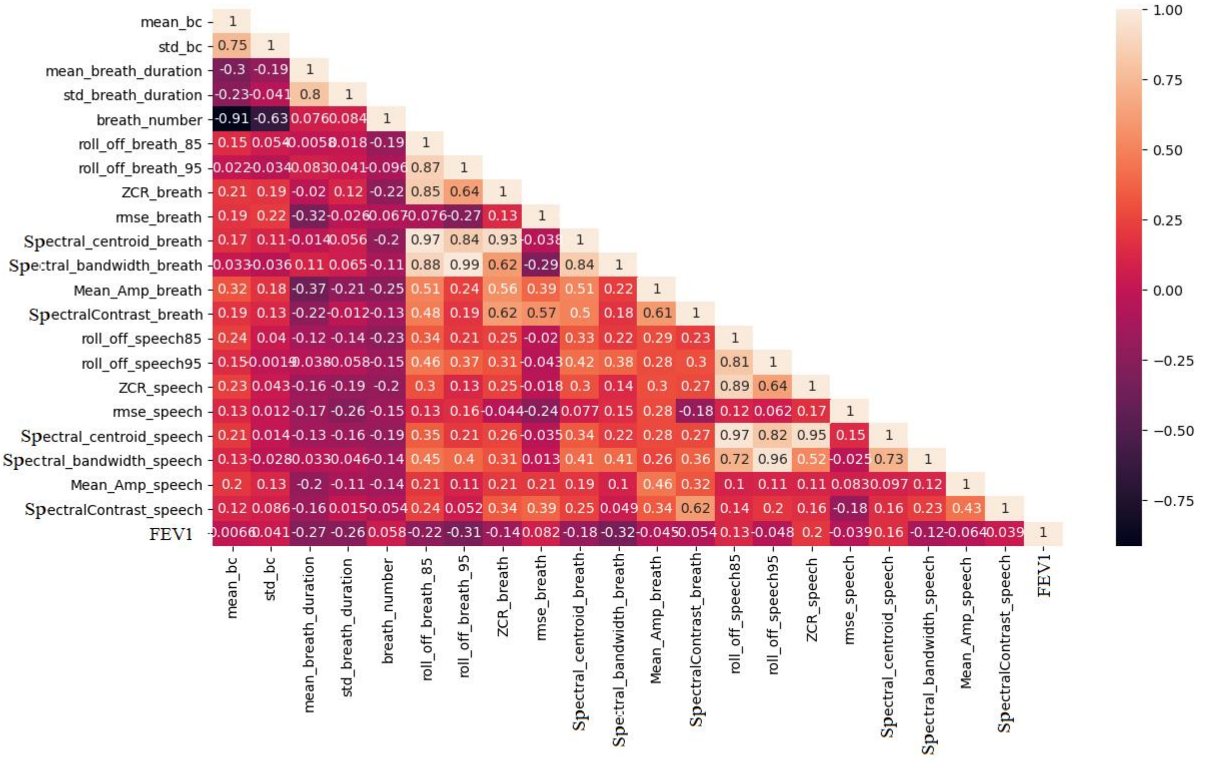


Supplementary Figure S3: Correlations of the features with each other. This figure represents the correlation of the features with each other. The Pearson correlation coefficient (PCC) has been calculated for each feature with others. Each box contains a correlation value between each other. The correlation values range from -1 to 1 where 1 means the features are highly correlated and -1 means they are also highly correlated but their correlation is negative.

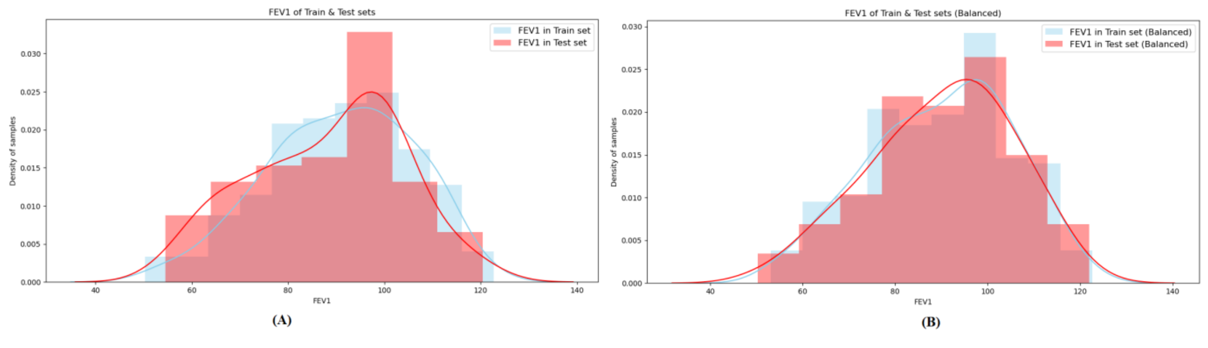


Supplementary Figure S4: The patterns of the training and the testing datasets. This figure represents the patterns of the training and the testing datasets. [1] the patterns of the training and the testing datasets without considering any special case. In this case samples are randomly separated for the training and testing sets at a ratio of 0.7:0.3 respectively. The distribution pattern of the samples on each FEV1% in the training and the testing sets show an small difference. [1] the patterns of the training and the testing datasets while the training and the testing samples are separately in a balancing way (please refer to the Section 4.1.4). In this case, samples are also randomly separated for the training and testing sets at the same ratio mentioned above. Additionally, the samples are separated at this ratio in each FEV1% range. The distribution pattern of the samples on each FEV1% range in the training and the testing sets present a similar pattern.

**Supplementary Tables:**

Supplementary Table S1: A threshold for separating the breath and speech chunks

| **Feature** | **Value** |
|---|---|
| Spectral contrast | 20.55 |
| Roll-off at 95% | 6000 |
| Root mean squared energy | 0.021 |
| Spectral bandwidth | 2300 |
| Mean amplitude | 0.018 |

Supplementary Table S2: American Thoracic Society Grades for Severity of a Pulmonary Function Test Abnormality ($FEV_1$% = forced expiratory volume in one second percentage predicted.

| Severity of lung function | $FEV_1$% |
|---|---|
| Normal | > 80 |
| Mildly abnormal | 70 to 79 |
| Moderately abnormal | 60 to 69 |
| Moderate to severely abnormal | 50 to 59 |
| Severely abnormal | 35 to 49 |
| Very severely abnormal | < 35 |

Supplementary Table S3: Features extracted from the voice sound files

| Macro Area | Feature |
|---|---|
| Breath Cycles Features | Mean breath cycle duration |
| | Breath cycle standard deviation |
| | Breaths number |
| Breaths Features | Mean breath duration |
| | Breath duration standard deviation |
| Breath and Speeches Features - Frequency domain | Roll off 85% |
| | Roll off 95% |
| | Zero Crossing Rate |
| | Root Mean Squared Energy |
| | Spectral Centroid |
| | Spectral Contrast |
| | Spectral Flatness |
| | Spectral bandwidth |
| | Mean Amplitude |

Supplementary Table S4: The performance in predicting normal vs. abnormal lung function resulted from the features from the voice features only ($Model_3$). Here, RF = Random Forest classifier, SVC = Support Vector Classifier and LR = Generalised Linear Regression method.

| | | **Class label** | **Precision** | **Recall** | **$F_1$-score** | **Accuracy** | **Sensitivity** | **Specificity** |
|---|---|---|---|---|---|---|---|---|
| RF | Abnormal | 0 | 0.75 | 0.44 | 0.56 | | | |
| | Normal | 1 | 0.81 | 0.94 | 0.87 | | | |
| | Macro average | | 0.78 | 0.69 | 0.72 | | | |
| | Weighted average | | 0.80 | 0.80 | 0.79 | | | |
| | Score | | 0.80 | 0.80 | 0.79 | 0.80 | 0.44 | 0.94 |
| SVC | Abnormal | 0 | 0.88 | 0.26 | 0.40 | | | |
| | Normal | 1 | 0.78 | 0.99 | 0.87 | | | |
| | Macro average | | 0.83 | 0.62 | 0.63 | | | |
| | Weighted average | | 0.80 | 0.78 | 0.74 | | | |
| | Score | | 0.80 | 0.78 | 0.74 | 0.78 | 0.26 | 0.99 |
| LR | Abnormal | 0 | 0.71 | 0.37 | 0.49 | | | |
| | Normal | 1 | 0.80 | 0.94 | 0.86 | | | |
| | Macro average | | 0.75 | 0.66 | 0.68 | | | |
| | Weighted average | | 0.77 | 0.78 | 0.76 | | | |
| | Score | | 0.77 | 0.78 | 0.76 | 0.78 | 0.37 | 0.94 |

Supplementary Table S5: The performance in predicting normal vs. abnormal lung function resulted from the voice features with sex, weight and height ($Model_{3P}$). Here, RF = Random Forest classifier, SVC = Support Vector Classifier and LR = Generalised Linear Regression method.

| | | Class label | Precision | Recall | $F_1$-score | Accuracy | Sensitivity | Specificity |
|---|---|---|---|---|---|---|---|---|
| RF | Abnormal | 0 | 0.77 | 0.63 | 0.69 | | | |
| | Normal | 1 | 0.87 | 0.93 | 0.90 | | | |
| | Macro average | | 0.82 | 0.78 | 0.80 | | | |
| | Weighted average | | 0.84 | 0.85 | 0.84 | | | |
| | Score | | 0.84 | 0.85 | 0.84 | 0.85 | 0.63 | 0.93 |
| SVC | Abnormal | 0 | 0.83 | 0.37 | 0.51 | | | |
| | Normal | 1 | 0.80 | 0.97 | 0.88 | | | |
| | Macro average | | 0.82 | 0.67 | 0.70 | | | |
| | Weighted average | | 0.81 | 0.80 | 0.78 | | | |
| | Score | | 0.81 | 0.80 | 0.78 | 0.80 | 0.37 | 0.97 |
| LR | Abnormal | 0 | 0.81 | 0.48 | 0.60 | | | |
| | Normal | 1 | 0.83 | 0.96 | 0.89 | | | |
| | Macro average | | 0.82 | 0.72 | 0.75 | | | |
| | Weighted average | | 0.82 | 0.82 | 0.81 | | | |
| | Score | | 0.82 | 0.82 | 0.81 | 0.83 | 0.48 | 0.96 |

### TRIPOD Checklist: Prediction Model Development

| Section/Topic | Item | Checklist Item | Page |
|---|---|---|---|
| **Title and abstract** | | | |
| Title | 1 | Identify the study as developing and/or validating a multivariable prediction model, the target population, and the outcome to be predicted. | 1 |
| Abstract | 2 | Provide a summary of objectives, study design, setting, participants, sample size, predictors, outcome, statistical analysis, results, and conclusions. | 2 |
| **Introduction** | | | |
| Background and objectives | 3a | Explain the medical context (including whether diagnostic or prognostic) and rationale for developing or validating the multivariable prediction model, including references to existing models. | 3-4 |
| | 3b | Specify the objectives, including whether the study describes the development or validation of the model or both. | 4 |
| **Methods** | | | |
| Source of data | 4a | Describe the study design or source of data (e.g., randomized trial, cohort, or registry data), separately for the development and validation data sets, if applicable. | 4 |
| | 4b | Specify the key study dates, including start of accrual; end of accrual; and, if applicable, end of follow-up. | 4 |
| Participants | 5a | Specify key elements of the study setting (e.g., primary care, secondary care, general population) including number and location of centres. | 4 |
| | 5b | Describe eligibility criteria for participants. | 4 |
| | 5c | Give details of treatments received, if relevant. | NA |
| Outcome | 6a | Clearly define the outcome that is predicted by the prediction model, including how and when assessed. | 5 |
| | 6b | Report any actions to blind assessment of the outcome to be predicted. | NA |
| Predictors | 7a | Clearly define all predictors used in developing or validating the multivariable prediction model, including how and when they were measured. | 4-6 |
| | 7b | Report any actions to blind assessment of predictors for the outcome and other predictors. | NA |
| Sample size | 8 | Explain how the study size was arrived at. | 4 |
| Missing data | 9 | Describe how missing data were handled (e.g., complete-case analysis, single imputation, multiple imputation) with details of any imputation method. | NA |
| Statistical analysis methods | 10a | Describe how predictors were handled in the analyses. | 4-5 |
| | 10b | Specify type of model, all model-building procedures (including any predictor selection), and method for internal validation. | 5-6 |
| | 10d | Specify all measures used to assess model performance and, if relevant, to compare multiple models. | |
| Risk groups | 11 | Provide details on how risk groups were created, if done. | NA |
| **Results** | | | |
| Participants | 13a | Describe the flow of participants through the study, including the number of participants with and without the outcome and, if applicable, a summary of the follow-up time. A diagram may be helpful. | NA |
| | 13b | Describe the characteristics of the participants (basic demographics, clinical features, available predictors), including the number of participants with missing data for predictors and outcome. | NA |
| Model development | 14a | Specify the number of participants and outcome events in each analysis. | 4 |
| | 14b | If done, report the unadjusted association between each candidate predictor and outcome. | NA |
| Model specification | 15a | Present the full prediction model to allow predictions for individuals (i.e., all regression coefficients, and model intercept or baseline survival at a given time point). | 6-8 |
| | 15b | Explain how to the use the prediction model. | 6-8 |
| Model performance | 16 | Report performance measures (with CIs) for the prediction model. | 7-8 |
| **Discussion** | | | |
| Limitations | 18 | Discuss any limitations of the study (such as nonrepresentative sample, few events per predictor, missing data). | 9 |
| Interpretation | 19b | Give an overall interpretation of the results, considering objectives, limitations, and results from similar studies, and other relevant evidence. | 9 |
| Implications | 20 | Discuss the potential clinical use of the model and implications for future research. | 10 |
| **Other information** | | | |
| Supplementary information | 21 | Provide information about the availability of supplementary resources, such as study protocol, Web calculator, and data sets. | 10 |
| Funding | 22 | Give the source of funding and the role of the funders for the present study. | 10 |

We recommend using the TRIPOD Checklist in conjunction with the TRIPOD Explanation and Elaboration document.
